# Robust image-based risk predictions from the deep learning of lung tumors in motion

**DOI:** 10.1101/2021.07.28.21261255

**Authors:** Amishi Bajaj, P. Troy Teo, James Randall, Bin Lou, Jainil Shah, Mahesh Gopalakrishnan, Ali Kamen, Mohamed E. Abazeed

**Affiliations:** 251 E. Huron St, Galter Pavilion LC-178, Department of Radiation Oncology, Northwestern University, Feinberg School of Medicine, Chicago, IL 60611; 755 College Road East, Digital Technology and Innovation Division, Siemens Healthineers, Princeton, NJ, 08540; 40 Liberty Blvd, Diagnostic Imaging Computed Tomography, Siemens Healthineers, Malvern, PA 19355; 303 E. Superior St/Lurie 7, Robert H. Lurie Cancer Center, Northwestern University, Chicago, IL 60611

## Abstract

**Introduction:** Deep learning (DL) models that use medical images to predict clinical outcomes are poised for clinical translation. For tumors that reside in organs that move, however, the impact of motion (*i.e*. degenerated object appearance or blur) on DL model accuracy remains unclear. We examine the impact of tumor motion on an image-based DL framework that predicts local failure risk after lung stereotactic body radiotherapy (SBRT).

**Methods:** An IRB-approved study (NU00212113) was used to identify patients treated with lung SBRT. Pre-therapy free breathing (FB) CT images from 849 patients were input into a multi-task deep neural network to generate an image fingerprint (or DL score) that predicts time-to-event local failure outcomes. The impact of tumor motion was examined on the DL scores for a hold-out set comprising 228 image series from 19 patients comprising: (1) FB CT, (2) four-dimensional (4D) CT, and (3) maximum-intensity projection (MIP) images.

**Findings:** Variance in image-derived DL scores were associated with tumor motion (*R*^2^ = 0.45). Specifically, DL score variance was deterministic with undulations in phase with the respiratory cycle (*R*^2^ = 0.77-0.99). DL scores, but not tumor volumes, peaked near end-exhalation (∼50% phase). The mean of the scores derived from 4D CT images and the score obtained from FB CT images were highly associated (Pearson ***r*** = 0.99). MIP-derived DL scores were significantly higher than 4D- or FB-derived risk scores (*p* < 0.0001).

**Interpretation:** We show that an image-based DL risk score derived from a series of four-dimensional CT images varies in a deterministic, sinusoidal trajectory in phase with the respiratory cycle. These results indicate that deep learning models of tumors in motion can be robust to fluctuations in object appearance due to movement and can guide standardization processes in the clinical translation of DL models for patients with lung cancer.

## INTRODUCTION

Stereotactic body radiotherapy (SBRT) enables the delivery of conformal radiation at very high doses resulting in improved local control at the irradiated site(s)^1^. Since its emergence, it has evolved to significantly alter the management of patients with early-stage lung cancers or oligo-metastatic disease^2-6^. Critically, increased utilization rates have occurred in conjunction with critical technical and clinical advancements that range from radiation delivery improvements (*e.g*. static to arc-based therapy fields^7^) to risk-adapted dose recommendations^4^. Despite these advances, there is an emergent category of studies that indicate that certain patients are more likely to experience local failure after SBRT^8-12^. In addition, inter-patient heterogeneity, *a priori*, suggests that uniform radiotherapy doses optimized for populations of patients can lead to the under- and over-treatment of a significant proportion of individuals^13^. Therefore, there is significant clinical utility in the identification and clinical implementation of determinants that can guide more personalized dose delivery.

We, and others, have identified several clinicopathological^9^, image^8^, and genetic^12^ features that can guide the use of individualized risk predictions for lung SBRT. Precision in the measurements of these variables at scale and across diverse clinical settings, however, is pivotal for the alignment of lung SBRT with clinically actionable personalized radiation dose treatment guidance. A critical step towards precision is to mitigate “noise” in variate measurements, which can be achieved by the adoption of algorithmic standards that amplify signals and dampen artifacts. Although measurements of genetic and clinical features can be subject to some variability^14,15^, high-dimensional datasets like image-based quantitative features can be particularly sensitive to fluctuations in the image source, acquisition parameters, and reconstruction algorithms^16-19^.

In addition to these potential sources of variation, tumor motion has been shown to impact radiomic feature stability. Lung tumors, in particular, can have significant motion during respiration^20^ and radiomic features extracted from free breathing (FB) computed tomography (CT)^21^ or four-dimensional (4D) CT scans can be affected by motion^22^. Tumor motion could be mitigated with active breathing control (breath hold) and use of abdominal compression techniques^12^. Despite limiting respiratory excursion, however, some degree of tumor motion is inevitable for the vast majority of patients, necessitating the accounting of motion’s effects on prediction model accuracy and precision.

Deep learning (DL) is the process of feeding a machine raw data and allowing it to discover vectors for classification through the use of multiple layers of features^23^. It holds significant, but currently unrealized, promise in clinical prediction and there has been very little progress in the use of DL to predict tumor responses to individual anticancer therapies^24^. We previously developed a deep neural network that creates new radiomic features via deformable multitasking to predict the risk of failure for patients treated with SBRT^8^. However, the impact of tumor motion on model accuracy was not evaluated. Critically, the extent that tumor motion affects the precision of DL diagnostic or theranostic (dose optimization) models remains unknown. Herein, we assess the impact of tumor motion in the output of a deep neural network for patients treated with lung SBRT.

## METHODS

### Study Population

A Northwestern University Institutional Review Board (IRB) approved study (NU00212113) was used to identify 283 patients treated with lung stereotactic body radiotherapy (SBRT) from 2010-2020. Patients with primary (stage IA-IV) or recurrent lung cancer as well as patients with other cancer types with solitary or oligometastases to the lung were included. Patients without digitally accessible CT (FB and 4D) image and radiotherapy structure data were excluded from the study. A total of 243 patients met our eligibility criteria, 19 patients were randomly selected for additional analyses (see below for randomization method).

Patients were treated based on either a pathological or radiographic diagnosis. All primary lung cancer patients were staged using a CT of the chest. PET and imaging of the brain (magnetic resonance imaging [MRI] or CT) was employed when clinically indicated. Patients were treated using custom immobilization devices. Abdominal compression to restrict breathing motion was at the discretion of the treating physician. The patient population used as input for the development (training and validation) of the DL model comprises 849 patients from nine distinct clinical sites who met the same eligibility criteria^8^.

### CT Image Dataset

CT images with corresponding physician-designated gross and internal tumor volumes (GTV or ITV) were used for our analysis. A Philips Brilliance CT Big Bore (Koninklijke Philips N.V.; Amsterdam, Netherlands) was used for the NU study population. FB CT images were obtained with a gantry rotation speed (∼1.5s) to produce a low-pitch (0.313) CT scan. All 4D CT images were acquired in cine with a 440 ms tube rotation time; the CT component is a 16-slice CT. A pressure sensor belt (Model AZ-733 V; Anzai Medical Co, Ltd; Tokyo, Japan) attached to the upper abdominal region was used as an external surrogate for organ motion, providing a respiratory trace of relative abdominal height over time. Cine 4DCT image sets were assembled using a low-pitch helical scanning technique where the pitch is set low enough that each slice of the volume remains illuminated for at least one duration of respiratory cycle. The detector collimation of 16 ⨯ 1.5 mm was employed which provided a 3 mm minimum slice thickness.

Acquisitions were obtained at 120 kVp, with a 100 mA tube current for clinical acquisitions, and a dose-sparing 50 mA for experimental acquisitions. Helical scans were processed retrospectively to yield a final 4DCT with a set of ten CT scans, each representing a component of the breathing phase labeled using the pressure sensor belt. A maximum-intensity projection (MIP) image, consisting of a pixel-by-pixel maximum CT number, was generated from 4D image set^25^. FB CT images were used to train the deep neural network and applied to a holdout set of FB, 4D, and MIP images in our current cohort.

### Deep Learning Score

All images were resampled to a 1.17 mm x 1.17 mm x 3 mm resolution. The original Hounsfield Unit (HU) values were clipped at -1,000 and 1000 and rescaled to -1 to 1 to better fit the input range of our model. The original CT volume for each patient was cropped to a 64 × 64 x 32 sub-volume or ∼7.5 cm x 7.5 cm x 9 cm, chosen to encompass all tumors to mitigate over-fitting, avoid spurious associations at a distance from the tumor, and incorporate putative tumor motion (the ITV). A physician delimited GTV mask was used as an additional channel rather than masking the CT volume, permitting flexibility for the network to learn features from both inside and outside of the GTV. Given that the image features extracted by the network is expected to be minimally influenced by the bounding box position (i.e. shift invariant) as long as the box encompasses the GTV, we randomly selected the cropped regions for each iteration of the training process. This data augmentation technique facilitated the network’s ability to locate the desired invariant.

A convolutional neural network (CNN) was utilized as an encoder for the extraction of imaging features; its use for image segmentation and classification and object detection has been well described.^15^ The input layer to the encoder network included 2 channels: CT image and ITV contour. The encoder consisted of 8 convolutional layers of 3 × 3 x 3 filter size. There were 16 filters in the first layer with the filter size doubled every two layers. There were max pooling operations after the second, fourth and sixth convolutional layers, with a pooling size of 2 × 2 x 2. The last convolutional layer was followed by an average pooling layer for extracting one feature from each feature map. Therefore, the encoder network reduced the feature dimension from 64 × 64 x 32 (input volume) to 128 (latent space). In all convolutional layers, we employed rectified linear units (ReLUs) as the nonlinear activation function. The CNN-based encoder network was used to extract image features and a fully-connected layer was adopted to combine all image features into one DL signature or score.

DL scores were then measured for FB CT, each respiratory phase in the 4D CT image sets, and on the maximum-intensity projection (MIP) scans.

### Tumor Motion

4D and FB CT images were superimposed using an image registration algorithm of the MIM Software package (MIM Software; Cleveland, OH). To estimate tumor motion in our current work, the GTV in the treatment plan was propagated to the other 10 breathing phases of the 4D CT images using Pinnacle Treatment Planning System (Philips Medical Systems, Fitchburg, WI). The 4D images and structures sets were then exported for analysis. A Matlab code (Mathworks, Inc) was used to extract the contours of the propagated GTV from the RTStruct DICOM header tags. The centroid of each GTV was determined by calculating the arithmetic mean position of all the coordinates defining the GTV. To quantify target motion, the 3D displacement (cm/phase) of the GTV centroid and the variations of the volume were computed. Distance traveled in each dimension (*x* = right-left, *y* = anterior-posterior, and *z* = superior-inferior motion) was summed across breathing phases and the root sum square [*i.e*. 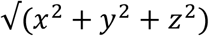], an aggregate measure of distances traversed in all dimensions, was calculated.

### Statistical Analysis

The “RAND” and “RANK” functions in Excel were used to randomly assign a deidentified code for each patient. Given α = 0.05 and 1-β (power) = 0.90, the required sample size to detect a deep-learning (DL) score association between two imaging datasets of magnitude *R*^2^ = 0.4 and corresponding effect size |***r***| or correlation ρ of H1= 0.63) is 19 patients. Therefore, the first 19 patients, from a numerically sorted list of de-identified codes, who met the inclusion criteria were selected for further analysis. Pearson’s correlation coefficient was used to measure the strength of associations between bivariates. Linear regression was used to assess the association between DL scores from FB and the mean DL scores obtained from each phase in the 4D image sets. One-way analysis of variance (ANOVA) was used to compare patients’ DL scores among FB, 4D, and MIP scans. A Wilcoxon matched-pairs signed rank test with a Geisser-Greenhouse correction was used to assess statistical significance. Sinusoidal regression was used to model variation of DL score as a function of respiratory phase on 4DCT for patients with low, intermediate, and high DL scores, with a determination coefficient generated for each of these models to quantify goodness of fit. Scatterplot curve fitting with 95% confidence interval projections of intra-patient normalized [smallest value = 0; largest value = 1] DL scores and volumes were calculated using locally estimated scatterplot smoothing (LOESS). *P* value of < 0.05 was considered statistically significant. Statistical analysis was performed using *R* (4.0.5; Vienna, Austria)^18^.

## RESULTS

### Patient Characteristics

From the 243 patients who met our eligibility criteria, 19 were randomly selected for additional analyses. 57.9% of the cohort had peripheral lung tumors, defined as tumors located 2 cm from the proximal bronchial tree and not in close proximity to critical organs like larger airways, heart, esophagus, and major arteries (Table 1). 47.4% of patients underwent abdominal compression to restrict breathing motion. The selected patients had treatments spanning a period of time (from 2012 to 2019) in which abdominal compression and CT simulation procedures were not substantively modified in our clinic. The median of aggregate motion in all dimensions was 1 cm [IQR: 0.72-1.53], indicating quantifiable tumor movement despite abdominal compression. Importantly, similar to the training dataset population, this cohort included patients with primary lung cancer as well as metastatic disease to the lung.

**Table 1.** Baseline Characteristics of the Study Population

| Characteristic | No. of Patients (%) or Median [IQR] |
| --- | --- |
| Age | 76.48 [65.35, 81.82] |
| Gender (Female/Male) | 13/6 (68.4/31.6) |
| Ethnicity (%) |  |
| African | 5 (26.3) |
| European | 11 (57.9) |
| Hispanic or Latino | 3 (15.8) |
| Year of treatment |  |
| 2012 | 1 ( 5.3) |
| 2015 | 3 (15.8) |
| 2016 | 4 (21.1) |
| 2017 | 1 ( 5.3) |
| 2018 | 7 (36.8) |
| 2019 | 3 (15.8) |
| Lobe |  |
| LUL | 7 (36.8) |
| RLL | 6 (31.6) |
| RML | 1 ( 5.3) |
| RUL | 5 (26.3) |
| Dose per fraction | 10.00 [10.00, 11.00] |
| No. of fractions | 5.00 [4.50, 5.00] |
| BED | 100.00 [100.00, 102.80] |
| Tumor location (%) |  |
| Peripheral | 11 (57.9) |
| Central | 8 (42.1) |
| Motion control | 9/10 (47.4/52.6) |
| Histology |  |
| Adenocarcinoma | 9 (47.4) |
| Clear Cell | 1 ( 5.3) |
| Clinical Diagnosis | 4 (21.1) |
| Squamous | 5 (26.3) |
| CT size (cm) | 1.70 [1.40, 2.50] |
| Overall stage |  |
| IA1 | 2 (10.5) |
| IA2 | 8 (42.1) |
| IA3 | 5 (26.3) |
| IIA | 1 ( 5.3) |
| IV | 3 (15.8) |
| PET SUVmax | 6.31 [3.15, 8.93] |
| Treatment intent |  |
| Curative | 14 (73.7) |
| Oligometastatic | 4 (21.1) |
| Salvage after Surgery | 1 ( 5.3) |
| Tumor motion (cm) | 1.00 [0.72, 1.53] |

Continuous variables are represented as median [IQR].
Abbreviations: BED, biologically effective dose; BMI, body mass index; IQR, inter-quartile range; PET, positron emission tomography; SUV, standardized uptake values.

### Deep learning scores vary in phase with tumor motion

We input 4D CT images into an independently validated deep neural network-derived prediction model^8^. Image scores from this model were previously shown to be associated with an increased risk of local failure after lung SBRT. Moreover, image-based DL scores provided complementary information to the clinical established variables of histological subtype and radiation dose^23^. First, we derived DL scores for each phase of the respiratory cycle (0%-90%). Kernel density estimates of these scores showed varied ranges, and correspondent standard deviations, across the cohort (**Fig. 1a**). We measured the summation of displacements (cm/phase) for each tumor centroid (total distance traveled) for each axis and calculated the root sum squared, or the Euclidean distance in three dimensions, as a surrogate of tumor motion. The standard deviations of the DL scores were significantly associated with tumor motion (*P* = 0.005; simple linear regression) (**Fig. 1b**). These results indicated that a significant proportion of the variation in patient DL scores obtained from distinct 4DCT scans were attributed to tumor motion.

**Figure 1.**
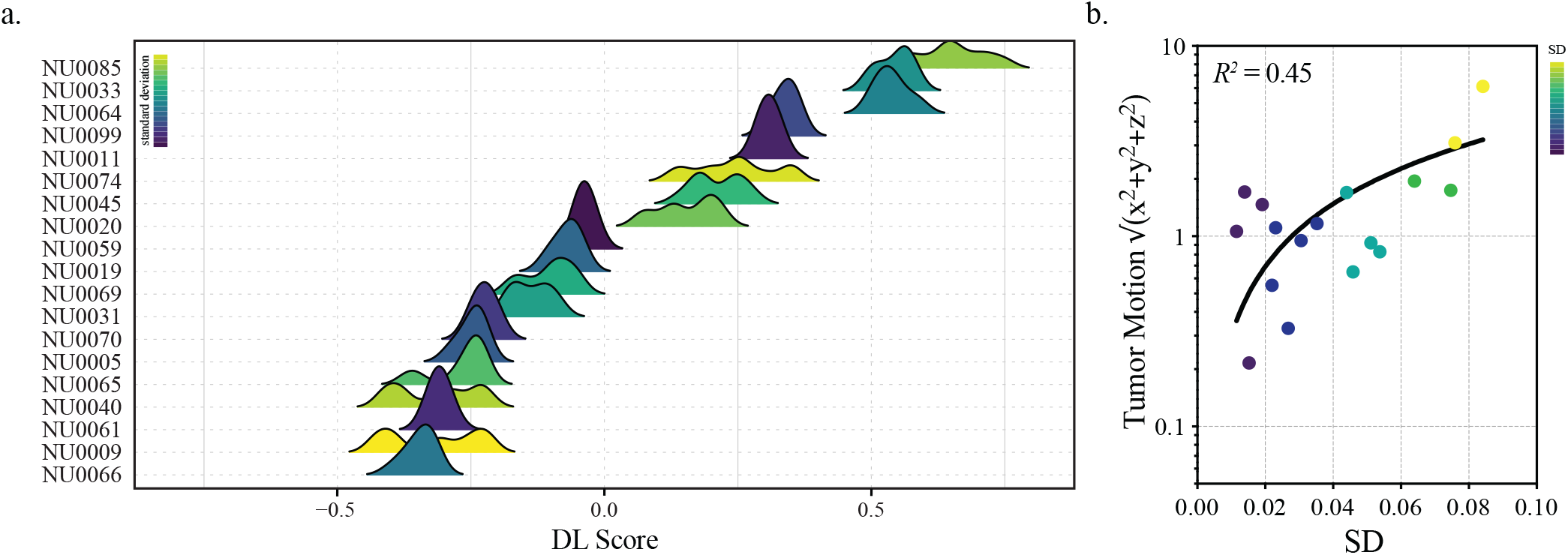
Deep learning score variation and tumor motion. (a) Probability density distribution plots of DL scores from ten 4DCT frames (0-90%). Plots are arranged by smallest to largest median score. (b) Association between tumor motion and standard deviation calculated in (a) are shown on a semi-*log* plot. Distance traveled in each dimension was summed across breathing phases and the root sum square for each patient was calculated. The coefficient of determination (*R*^2^) is based on linear regression.

Although tumor motion conferred fluctuations in DL scores, it remained unclear whether the observed variance was stochastic or deterministic. The former would entail a random and the latter a predictable variance as a function of object motion. To assess the relationship between DL scores, tumor motion, and the respiratory cycle in greater detail, we first sought precise measurements of tumor trajectories during the respiratory cycle. Although phase-binning approaches typically associate the inhale peak at 0% and the exhale peak at 40-60%, phase angle sorting is not consistent across patients^26^. Therefore, we verified end-inhale and end-exhale classifications, re-binning where appropriate, across our cohort using tumor centroid tracking and visual heuristics (**Fig. 2a**,**b**). A directed analysis of DL scores as a function of respiratory phase for select “mobile” tumors with high, intermediate, and low scores demonstrated striking sinusoidal trajectories (Fig. 2c).

**Figure 2.**
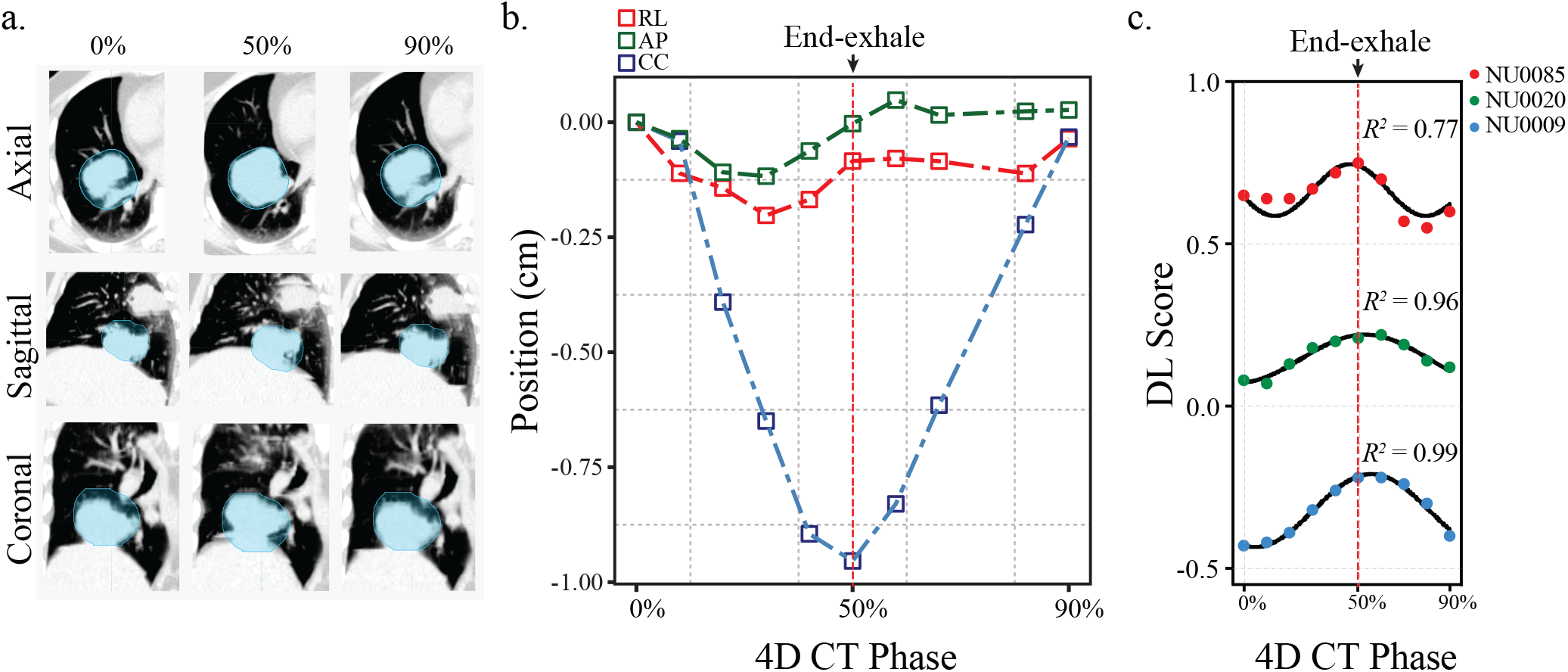
Tumor motion and the respiratory cycle. (a) Representative CT images at 0%, 50%, and 90% phases in three planes. Based on the binning algorithm and visual heuristics, end-inhalation = 0% and 90% and end-expiration = ∼50%. (b) The correspondent trajectory of the tumor centroids in (a) in 3D along left–right (LR), anterior–posterior (AP), and cranial–caudal (CC) directions across respiratory phases. (c) DL scores selected to represent high, intermediate, and low scores were fitted to a sine wave with a nonzero baseline.

To assess these trends across the cohort, we normalized the DL scores within patient and plotted these values as a function of respiratory phase (**Fig. 3a**). These results indicated that DL scores were deterministically higher near end expiration, returning to baseline at end inhalation. To determine if the changes in DL scores at end-expiration were tumor volume-dependent, we measured GTV across the respiratory cycles for each patient, normalized these values, and associated them with respiratory phase (**Fig. 3b**). In contrast to DL scores, there were no discernible changes in GTV across the respiratory cycle in our cohort. These data indicate that DL score vary in a deterministic, sinusoidal trajectory in phase with the respiratory cycle.

**Figure 3.**
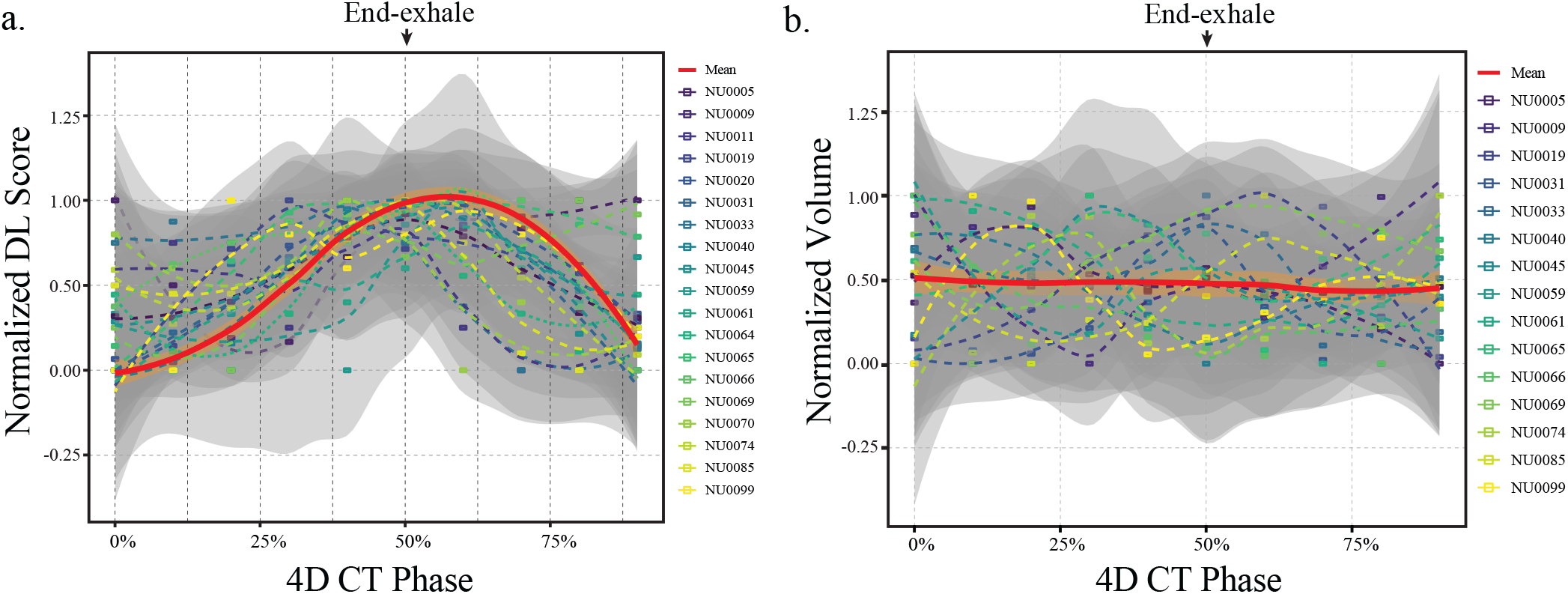
Deep learning scores peak at end-expiration. Intra-patient normalized scores (a) and tumor volumes (b) are plotted as a function of respiratory phase. Shaded gray represents the 95% CI of the locally estimated scatterplot smoothing fit for individual patients. Shaded orange represents the 95% CI of the of the locally estimated scatterplot smoothing fit of the mean values of the cohort per phase. Normalization [the smallest value = 0; the largest value = 1].

### DL scores correlate across distinct image types

To assess the associations between DL scores and image type, we combined the scores obtained from FB, 4D, or MIP scans from our cohort and measured the extent of local and global correlation in a matrix (**Fig. 4a**). The range of the correlation coefficients was high across all image types including FB, 4D phases, and MIP scans (Pearson r = 0.971-0.998). The means of the DL scores from the 10 distinct frames of the 4D image set were most associated with the end-inspiration scans, while the DL scores from the FB image set were most associated with end-exhalation scans. Critically, the means of the DL scores from the 10 distinct frames of the 4D image sets were highly associated with the DL scores obtained from the FB scans (Pearson *r* = 0.99; *P* < 0.0001) (**Fig. 4b**). The lowest correlation was noted between the scores of the MIP and end-inhale scans (0-20% or 90%) (**Fig 4a**). Correspondingly, the DL scores on the MIP scans were significantly higher than the DL scores obtained from the FB (*P* < 0.0001) or the 4D CT scans (*P* = < 0.0001). Altogether, there results indicated a high correlation between DL scores obtained from FB and 4D images. MIP scans, although retaining some associations with DL scores from other scan types, had distinctly higher values across the cohort.

**Figure 4.**
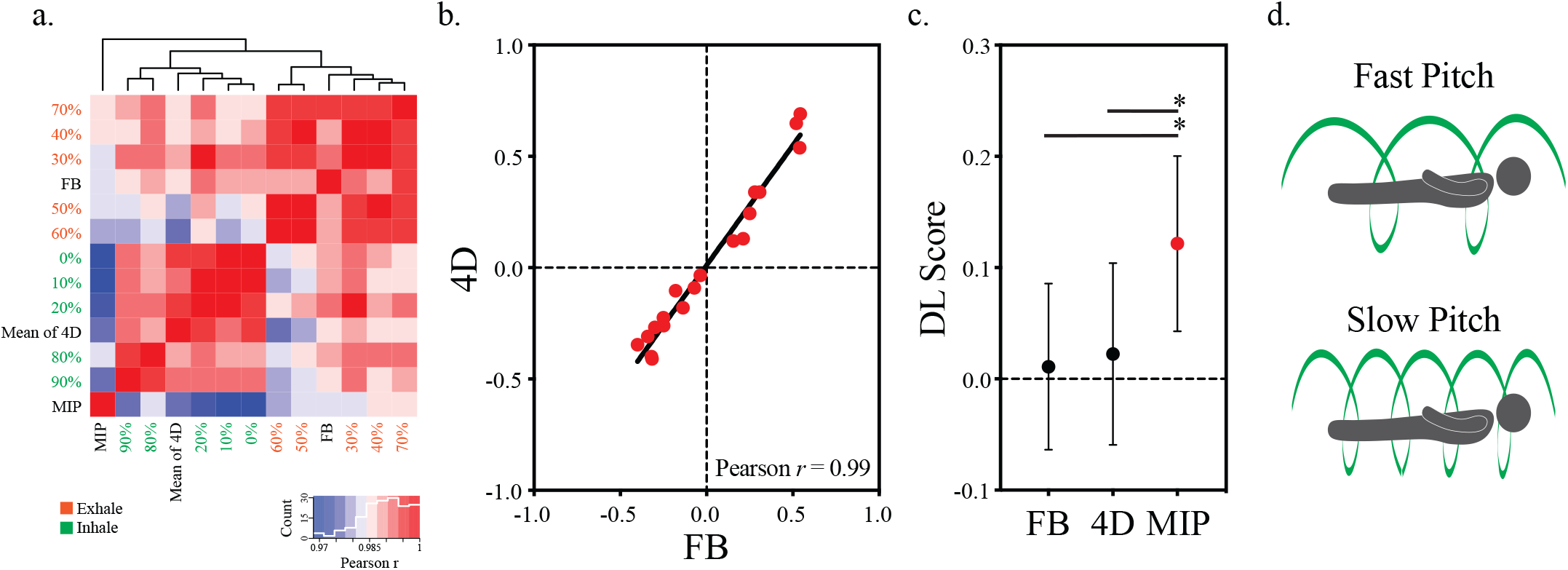
Deep learning score are highly associated across distinct image types. (a) Correlation matrix heatmap and dendrogram representing associations among DL scores and image types. The length of the dendrogram branches represents the distance between variables or clusters of variables calculated from bivariate Pearson correlations. (b) DL scores from FB and 4D CT scans were compared by scatterplot and linear regression (Pearson). Data points represent individual scores. (c) The mean ± S.E.M are plotted for the designated image categories. A Wilcoxon matched-pairs signed rank test was used to compare the groups (\**P* < 0.0001). (d) Schematic depicting the helical configuration of the beam traversing the patient at “fast” or “slow” table speed. Fast pitch scans are more likely to capture the tumor in a particular phase of the respiratory cycle.

## DISCUSSION

We previously developed a multi-task deep neural network to generate an image fingerprint that predicts time-to-event treatment outcomes after lung SBRT^8^. The output from the model was used to project an individualized dose of radiation that ameliorates the risk of local failure after SBRT to <5%. Although the model was independently validated using a plausibly related external study population, a source of variation that has yet to be accounted for in this, and similar lung-based DL models, is tumor motion. In this study, we show that a model trained using FB chest CT images undulates deterministically as a function of tumor respiratory excursion. The most important message in our study is that despite predictable fluctuations in the outputs, variations introduced by tumor motion do not substantively alter clinically actionable projections.

Previous work has shown that tumor motion can alter the radiomic feature space in discrete ways^21,22,27^. However, whether motion-associated variations are deterministic, capturing true changes in tumor composition, or stochastic, representing “noise” to be dampened remained unknown. Here, we show that DL scores varied in a predictable sinusoidal trajectory in phase with respiration. Intriguingly, the DL scores peaked at end-expiration and receded during inhalation across the cohort, and these changes were not associated with changes in tumor volumes. Since DL scores are trained on the primary outcome of local failure, these results suggest that tumors are marginally more sensitive to treatment in the deep inspiration phase of a respiratory cycle. Since most retrospective series are not sufficiently powered to detect such marginal differences, it remains unclear if deep inspiration breath-hold treatments could demonstrate a small improvement in local control outcomes.

Although the documented instability in the classical radiomic feature space created by tumor motion has been established^21,22,27^, it remains unclear if these variations are clinically meaningful. Moreover, motion-related variations in some radiomic features may not be relevant to DL models. Although some DL models can be fragile to small perturbations in input data^28-30^, our results indicate that a model built using FB scans and applied to tumors across distinct image types is not particularly fragile. These data indicate that DL models are not inherently prone to erroneous predictions when presented with distinct image types of the same tumor, even when tumor configuration is altered because of motion.

DL scores derived from MIP images, which represent a projection of voxels with the highest attenuation value on each imaging slice^16,17^, were systemically higher in our cohort. Despite these differences, MIP scores remained correlated with scores obtained from other scans from the same patients, although to a lesser extent than other image types. These data caution against the admixture of distinct image sets for training, validation, and testing, since systemic errors can affect projection accuracies. DL models poised for clinical testing should therefore be subject to rigorous standardization of image acquisition protocols and quality assurance testing before deployment. Measure of robustness, including adversarial image challenges, may also be warranted prior to implementation. Although recent studies indicating a tradeoff between robustness and accuracy should be considered^31^.

Our clinical standard for lung SBRT planning is the utilization of a low or “slow” pitch FB scan (**Fig. 4d**). This mode of image acquisition allows for an improved representation of the feature space of the tumor in motion compared to individual frames derived from the 4D images, which are obtained using a fast helical pitch^24^. Accordingly, adopting this standard may lower variations that may arise during fast image acquisitions that may incidentally capture tumors at the extremes of the respiratory cycle. In addition to maintaining consistency with our historical dataset, adopting this standard also provides complementary image types that can be used to enhance model training. The use of other image types, including 4D images and their reconstructions (e.g. MIP or average intensity projections), in the training process of our network has yet to be explored.

The strengths of our study include the grouped nature of the data (multiple scans from the same patient), the use of image-based index score as a backbone for our analyses, and the independent means of capturing tumor motion (*i.e*. tumor centroid approximations). The limitations of our study include the following: (1) we cannot fully account for all potential causes of bias; (2) the test cohort, despite being powered to detect the demonstrated effect size, is limited in size; and (3) we do not assess the impact of tumor motion on model training. These limitations can, in part, be addressed with the incorporation of additional cases and new image sets that incorporate distinct phases of respiration into model training and validation.

In conclusion, this is the first study to assess the impact of tumor motion on the output of a deep neural network. These findings indicate that tumor motion can yield deterministic changes in the output of the DL model and therefore should not be viewed reductively as artifact to be suppressed. Strict standardization of CT image use for model training, validation, and testing prior to clinical applications is advised to reduce the possibility for the propagation of systemic error.

## Data Availability

The datasets analyzed during the current study will be available from the corresponding author at the time of publication. Per institutional policy, the datasets are designated limited access. Upon receiving access, the investigator may only use them for the purposes outlined in the request to the data provider and redistribution of the data is prohibited.

## ACKNOWLEDGEMENTS

M.E.A. was supported by NIH R37CA222294, NIH P30CA060553, and the American Lung Association.

## CONTRIBUTORS

A.B., P.T., and J.R. assisted with data generation and analysis. A.B. contributed to the writing of the manuscript. B.L., J.S., A.K., and M.G. assisted with interpretation and edited the manuscript. M.E.A. conceived, designed, analyzed, interpreted, and supervised the overall work and wrote the manuscript.

## DECLARATION OF INTERESTS

B.L., A.K., and M.E.A. are named inventors in a patent for the use of Deep Profiler and *i*Gray to personalize radiotherapy dose. The concepts and information presented here are based on research results that are not commercially available. Future commercial availability cannot be guaranteed.

## Notes

### Competing Interest Statement

B.L., A.K., and M.E.A. are named inventors in a patent for the use of Deep Profiler and iGray to personalize radiotherapy dose.

### Author Declarations

A Northwestern University Institutional Review Board (IRB) approved study (NU00212113) was used to identify 283 patients treated with lung stereotactic body radiotherapy (SBRT) from 2010-2020.

